# The Effective Coverage of Maternal and Child Primary Health Care Services and its Relationship with Health Expenditures: An Analysis at Sub-National Level in Iran

**DOI:** 10.1101/2021.09.01.21262767

**Authors:** Elham Abdalmaleki, Zhaleh Abdi, Saharnaz Sazgarnejad, Bahar haghdoost, Elham Ahmadnezhad

## Abstract

**Backgrounds:** Measuring the effective coverage of essential health services is necessary for monitoring progress towards Universal Health Coverage (UHC). So, this study aimed to assess the geographic variations in key maternal and child indicators (as essential health services) provided at the primary health care (PHC) level in terms of their crude and effective coverage, and also to investigate the relationship between the effective coverage and health expenditures in the national and sub-national level of Iran.

**Methods:** This study was a secondary analysis, which analyzed the spatial distribution of six key maternal and child health indicators using the latest available data of Demographic Health Survey-DHS (2010) across 31 provinces of Iran. Moreover, two composite indicators, the crude, and the effective coverage were calculated. The median cut-off was used to compare provinces’ situations. Furthermore, the relationship between coverage indicators and total health expenditure per capita was evaluated.

**Results:** At the national level, the crude and the effective composite coverage were 89.56 and 77.22%, respectively. Also, the medians of composite crude and effective service coverage in the provinces were 90.25 and 77.62%, respectively. There was no significant difference between urban and rural areas.

**Conclusions:** in this study, we found that there is a significant gap between crude and effective service coverage of the selected indicators. Overall, coverage indicators of maternal services were higher compared to those of children. In addition, geographic variations in key Indicators of maternal and child health services coverage among provinces were almost high. Although the services are free of charge in the rural areas, they did not have higher coverage than those of urban areas. PHC services in Iran are far away from reaching the desired coverage and achieving UHC.

What is already known?

- In Iran, primary health coverage(PHC) has a long history, even years before the adoption of Alma-ata declaration in1979.
- Iran is committed to implementing PHC programs in line with the Astana Declaration to accelerate access to universal health coverage.

What are the new findings?

- there is a significant gap between crude and effective coverage of maternal and child services, with high geographic variations among provenances.
- Health expenditures have no statistical relationship between coverage indicators; however, in low-coverage provinces, per capita health expenditure was lower

What do the new findings imply?

- The concept of the effective coverage of interventions has not received enough attention from health policy-makers in Iran, so it has not been incorporated into health system performance, yet.
- Despite the long history of PHC in Iran, these services are still far away from reaching the appropriate targeted coverage; therefore, it is necessary to formulate and implement appropriate interventions in the provinces, based on their health needs and priorities.

## Introduction

### Primary Health Care services in the world and Iran

In 1978, the Alma-Ata Declaration defined the concept “primary health care (PHC)” as a strategy and a set of activities to provide equitable and affordable health services for reaching the goal of “health for all by the year 2000”[1]. In this regard, many countries have tried to reorganize their own health system based on PHC values over the past forty years. Notably, PHC approach has been known as a cornerstone of achieving global goals in Universal Health Coverage (UHC) and the health-related Sustainable Development Goals (SDGs). The Astana Declaration in 2018, 40 years after Alma Ata, was passed with the aims of strengthening PHC and accelerating progress towards the Sustainable Development Goals[2, 3]. Correspondingly, during the past decades, Iran has attained remarkable achievements in addressing nationwide inequity as well as the provision of primary health services to its rural areas[4].

### PHC transitional periods in Iran

PHC in Iran has experienced five rounds as follows: The first 12-year-round (1972-1983) was considered as the initial round that was for the development of the program[5]. During this period, the School of Public Health of Tehran University of Medical Sciences and the Ministry of Health in collaboration with the World Health Organization (WHO) implemented a study in “Rezaiyeh” (Currently named Urmia), which is known as the Rezaiyeh Study[6]. Accordingly, it led to the establishment of health houses to deliver PHC services in rural areas. In the second 12-year-round (1984-1995); the health care networks, health houses, and Behvarz training centers were developed in all regions of the country, which achieved international recognition. In the third 12-year-round (1996-2005), the selection and the grade of the Behvarzes’ education were revised, and the Rural Family Physician Program and Rural Citizens Health Insurance Program were introduced and then started in 2005[7]. The implementation of the family physician program in rural areas, to some extent, has led to setting up the health referral system for the rural population. However, *the referral system has not* been implemented successfully in urban areas and is only respected at the primary care level in rural areas[5,8]. In the fourth 12-year-round (2006-2018), the most important actions were taken in the form of a series of comprehensive health reforms in the health system, which is called Health Transformation Plan(HTP) with the main goal of achieving UHC by 2014[9]. Achieving UHC has been set as one of the targets of most of the countries worldwide while adopting the SDGs[10]. Also, the interventions to strengthen PHC services in Iran included earlier programs revision and the establishment of new advanced programs by focusing on integrating essential *non-communicable diseases (NCDs) services* into the *PHC* setting. Correspondingly, improving public health interventions, empowering health workforces, establishing PHC health facilities in urban and suburban areas for the first time, improving the electronic data recording system, and doing some activities to improve public health literacy were among the most important actions in this period[9].Finally, the fifth round in Iran has been started with the declaration of the Astana for PHC in 2019[3]. Iran, similar to other countries, is committed to implement the Astana declaration.

The significant reduction in maternal and neonatal mortality and the high coverage of universal vaccination are among the achievements of the PHC program in the above-mentioned rounds in Iran[11]. Moreover, in recent years, UHC has been at the center of the global public health agenda, and monitoring UHC is a part of the regular system of health systems performance assessment worldwide[12].

### PHC main measurements and target groups

It has been widely recognized that service coverage dimension of UHC should consider the quality of health services. This means that, in contrast to crude coverage that solely focuses on intervention access or usage, effective coverage is a measure that unites intervention need, use, and quality. The crude coverage indicator shows whether people have access to the services and utilize them or not. The effective coverage shows whether the provided health services are in line with the real demands of the targeted population. Moreover, are these people satisfied with receiving these services? Therefore, the outcome of PHC services can be measured with the mentioned indicators.

Two important groups that have been considered in all PHC rounds in Iran are mothers and children aged less than five years old[4]. The health related indicators of them, as the most important groups, are currently monitored and evaluated in the third goal of SDGs (and UHC).

Following the introduction of the SDGs and UHC, a general tendency has been formed to use composite indicators to compare the results obtained from the health interventions. For instance, in the world UHC reports, an indicator, called the Composite Coverage Index (CCI), has been used to compare the coverage of health services among countries[10,13]. In this study, to measure and compare PHC system performance among different provinces of Iran, composite indicators for both crude and effective coverage of a number of indicators were measured.

Another critical issue in achieving UHC is total health expenditure (THE) per capita, which has recently received much attention. It is noteworthy that actions taken in the health system including the PHC sector can be affected by the amount of funds allocated[13,14]. In this study, by assuming that provinces with higher health expenditure per capita may have better health outcomes, its impact on crude and effective coverage was assessed at the sub-national level. Therefore, this study aimed to assess the sub-national variations in key maternal and child indicators provided at PHC level in terms of their crude and effective coverage across the country, as well as investigating their relationship with health expenditures across different parts of the country.

## Methods

This study was a secondary analysis of the latest data of Demographic and Health Survey (DHS-2010), which is a combination of DHS-6 and Multiple Indicators Survey (MICS)-4[15]. The sample size was consisted of 31,350 households[16]. The obtained data were analyzed at both national and provincial levels (in 31 provinces), in terms of place of residence (urban and rural areas) and gender (for children indicators). Crude and effective coverage were also calculated for each indicator separately. In addition, composite crude and composite effective coverage was calculated for each category of indicators (i.e. maternal indicators and children indicators). Finally, the relationship of these two composite indicators with health expenditure per capita was assessed at the sub-national level.

### Indicators of crude and effective coverage for maternal and child primary health care services

In this study, we selected a range of key maternal and child health services to study the geographical distribution of health service utilization at district level. The indicators and the definition of their crude and effective coverage are presented in Table 1[17].

**Table 1:**
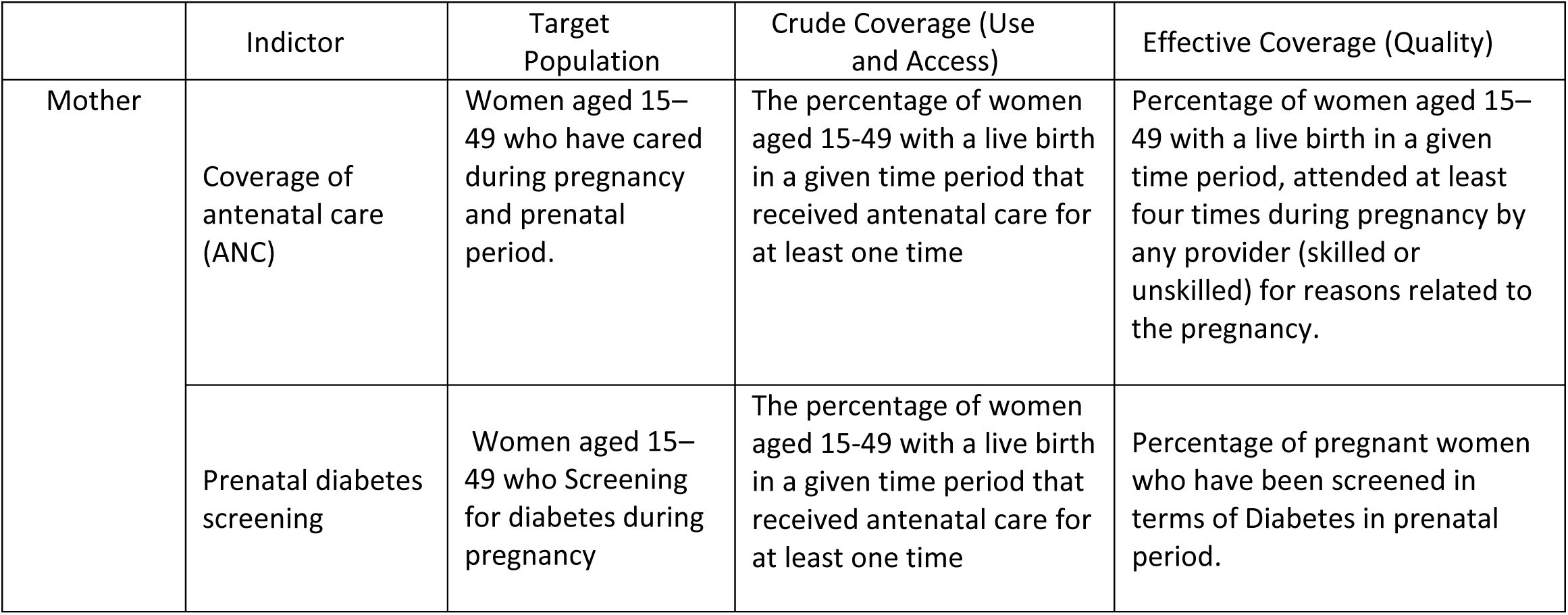
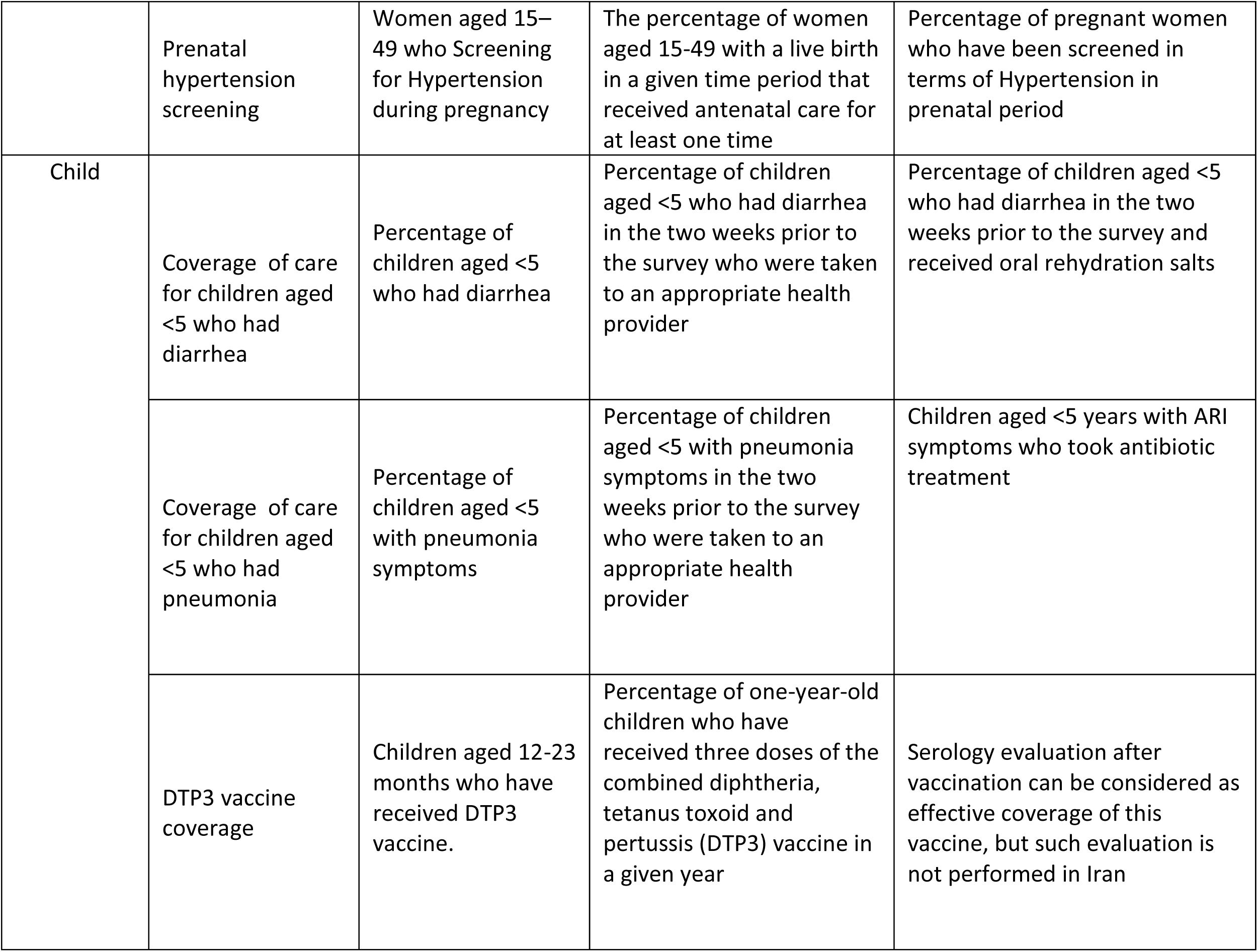
Definition of selected indicators, target population, Crude Coverage (Use) and Effective Coverage (quality) of services.

|  | Indictor | Target Population | Crude Coverage (Use and Access) | Effective Coverage (Quality) |
| --- | --- | --- | --- | --- |
| Mother | Coverage of antenatal care (ANC) | Women aged 15–49 who have cared during pregnancy and prenatal period. | The percentage of women aged 15-49 with a live birth in a given time period that received antenatal care for at least one time | Percentage of women aged 15–49 with a live birth in a given time period, attended at least four times during pregnancy by any provider (skilled or unskilled) for reasons related to the pregnancy. |
|  | Prenatal diabetes screening | Women aged 15–49 who Screening for diabetes during pregnancy | The percentage of women aged 15-49 with a live birth in a given time period that received antenatal care for at least one time | Percentage of pregnant women who have been screened in terms of Diabetes in prenatal period. |
|  | Prenatal hypertension screening | Women aged 15–49 who Screening for Hypertension during pregnancy | The percentage of women aged 15-49 with a live birth in a given time period that received antenatal care for at least one time | Percentage of pregnant women who have been screened in terms of Hypertension in prenatal period |
| Child | Coverage of care for children aged <5 who had diarrhea | Percentage of children aged <5 who had diarrhea | Percentage of children aged <5 who had diarrhea in the two weeks prior to the survey who were taken to an appropriate health provider | Percentage of children aged <5 who had diarrhea in the two weeks prior to the survey and received oral rehydration salts |
|  | Coverage of care for children aged <5 who had pneumonia | Percentage of children aged <5 with pneumonia symptoms | Percentage of children aged <5 with pneumonia symptoms in the two weeks prior to the survey who were taken to an appropriate health provider | Children aged <5 years with ARI symptoms who took antibiotic treatment |
|  | DTP3 vaccine coverage | Children aged 12-23 months who have received DTP3 vaccine. | Percentage of one-year-old children who have received three doses of the combined diphtheria, tetanus toxoid and pertussis (DTP3) vaccine in a given year | Serology evaluation after vaccination can be considered as effective coverage of this vaccine, but such evaluation is not performed in Iran |

### Composite Coverage Index-CCIs (Crude and Effective)

The composite coverage index was the weighted average coverage of six preventive and curative interventions received along with the continuum of maternal and child care. Accordingly, these interventions are as follows: (i) antenatal care coverage (ANC) visits by a skilled provider; (ii & iii) screening for pregnancy diabetes and hypertension; iv) oral rehydration therapy among children under five years old with diarrhea; *v)* suspected acute respiratory infection *care seeking among children under five years old; and vi)* Diphtheria-Pertussis-Tetanus (DPT) 3-three doses for the mentioned diseases immunization. Moreover, the following formula was used to calculate composite crude and composite effective coverage indicators[18]:

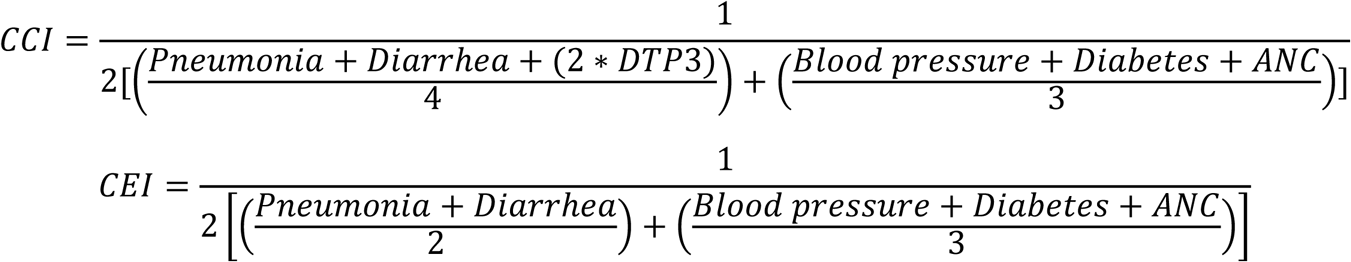

Composite Coverage Index gives weight equal to four stages in the continuum of care except for coverage of DTP3 immunization, which receives a weight of two, because more than one dose of the vaccine is required[18].

#### Relationship between crude and effective coverage of the indicators at the provincial level

Pearson correlation was used to examine the relationship between crude and effective coverage at the provincial levels. Afterward, a scatter plot was drawn to show this relationship. By considering a number of cut-offs, the chart was divided into four zones to examine geographic variations among provinces. Correspondingly, the first, second (median), and the third quartiles were calculated. Notably, out of them, the second (median) quartile is shown with a solid line on the scatter plot as the main cut-off. The first and third quartiles are also shown on the scatter plot with the dash-line as alternative cut-offs, to represent the exact performance of the provinces based on these indicators. In addition, the provincial map of the crude and effective coverage is illustrated based on the same cut-off points (the first to the third quartile). The scatter plot was drawn using R-software, and the ArcGIS software was used to draw the map.

### THE per capita and its effect on composite effective coverage

To compare the relationship between THE per capita and composite effective coverage, THE per capita in 2010 was extracted for each province[19]. The relationship is shown on the graph using bubble plot, linear regression fit model, and confidence interval considering the population weight. The relationship between THE per capita and composite effective coverage was also investigated using plotting and fitting linear regression fit model.

### Patient and Public Involvement

Study participants or the public were not involved in the design, or conduct, or reporting, or dissemination plans of our research.

## Results

The results are presented in four sections as follows: 1. Crude and effective coverage of the selected indicators for maternal and child care; 2. Composite indicators of crude and effective coverage at the national, provincial, urban, and rural levels; 3. Provincial distribution of the crude and effective indicators; and 4-the relationship between the composite effective coverage and THE.

### A Crude and effective coverage indicators by province, gender, and place of residence

crude and effective coverage of the main indicators in terms of the province, gender, and place of residence (rural/urban), which provided in the supplementary materials, varies from 33.29% for receiving care for children with diarrhea in some provinces to 100% for ANC, DTP3 vaccination, and screening for high blood pressure and gestational diabetes in pregnant women.

Also, among these indicators differences between rural and urban areas as well as differences between crude and effective coverage have been shown.

### B Composite Coverage index

#### i The National level

In 2010, at the national level, composite crude coverage and composite effective coverage averages were obtained as 89.56% and 77.2% for all indicators, respectively. The lowest amounts of crude and effective coverage were observed in Sistan-Baluchestan province (81.6% and 63.6%, respectively). Also, the lowest crude coverage was for diarrhea treatment in Markazi province with 38.4%, and the lowest effective coverage for the same indicator was for Kermanshah province with 33.3%. Maternal care indicators had the highest rates of crude and effective composite coverage (97.2% and 92.5%, respectively). For DTP3 vaccination indicator, only the crude service coverage was provided among the provinces, rather than the effective coverage, since information on the quality of the provided interventions was not available.

#### ii The subnational level

The provincial distribution of the indicators in 2010 is shown in figure 1, which indicates that they have no uniform distribution. As shown, the provinces with high crude coverage did not necessarily had a high effective coverage. In four provinces (Semnan, Bushehr, Lorestan, and Chaharmahal Bakhtiari), PHC interventions had both sufficient coverage and were effectively provided. Sistan-Baluchestan province had the poorest crude and effective coverage.

**Figure 1.**
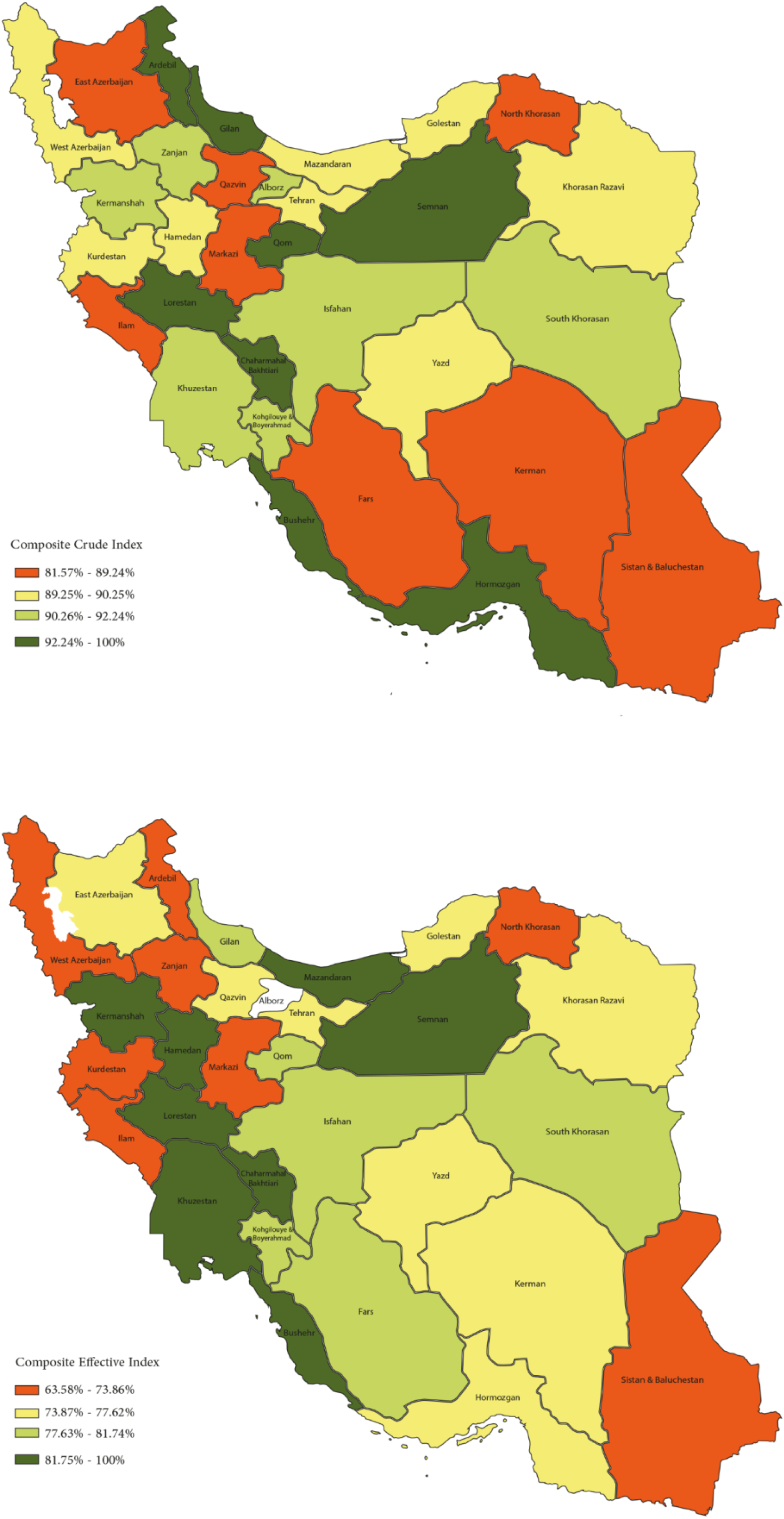
Composite crude and effective Coverage of primary health care services (six Interventions) in 31 provinces.

#### iii The urban and rural level

Crude coverage was 90.2% in urban areas compared to to 88.6% in rural areas. Effective coverage across rural and urban areas was 79.3% in urban areas in contrast to 73.9% in rural areas. Based on the analysis, coverage in urban areas is slightly higher than those of in rural areas.

### C The performance assessment of the provinces in PHC based on crude and effective coverage providing maternal and children services

Figure 2 plots composite crude service coverage against composite effective coverage or quality of care for each province, to identify health system priorities. The median values of crude and effective service coverage were 90.2 and 77.6%, respectively. In this regard, these median values were selected as the main cut-off points, and they were shown on the chart with the solid line. By applying the cut-offs, the figure was divided into four zones, and the provinces located in the lower-left zone were identified as the lowest performing provinces and also the provinces located in the upper right zone were identified as the highest performing provinces in the provision of maternal and child PHC services.

**Figure 2:**
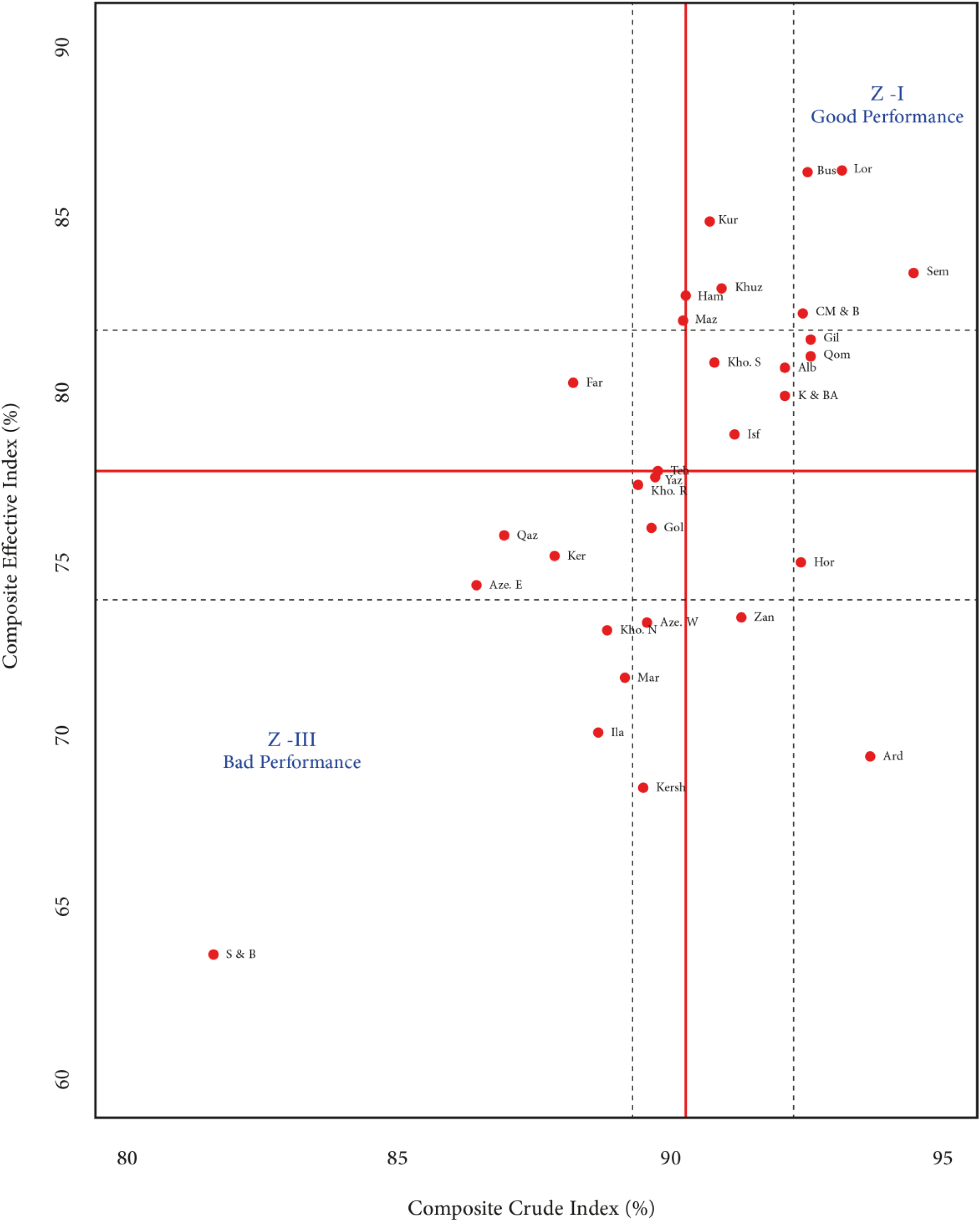
Composite effective coverage index against composite crude coverage index by provinces in 2010.

The dash-line on the scatter plot displays the performance of the provinces using the first and third quartiles, which were 89.2 and 92.2% for crude coverage, and 73.9 and 81.7% for effective coverage, respectively. According to the proposed cut-offs, Sistan-Baluchestan province had the lowest values of both crude and effective coverage of services. The provinces of Lorestan, Bushehr, Semnan, and Gilan have acceptable levels for both crude and effective coverage.

### D. The relationship between composite indicators and THE

The relationship between THE per capita and composite effective coverage (considering the population weight) in 2010 is graphically illustrated in figure 3. As shown, Sistan-Baluchestan province had the lowest THE per capita and a low coverage of services provision. Fars province had the highest per capita as well as an appropriate level of coverage, which indicates a direct relationship between expenditure per capita and service coverage. However, this direct relationship was not seen for Alborz province, since this province had relatively good coverage with a low per capita THE (It should be noted that Alborz province was formed by dividing Tehran province into two provinces, after obtaining the parliamentary approval in June 2010. Most likely, in 2010, all or some parts of its health funds were still provided from health funds of Tehran province. In other words, Alborz could be recognized as an outlier that should not be interpreted separately). To better understand the relationship between THE per capita and the service coverage, a linear regression fit model was performed. Accordingly, using the linear regression fit model, it was shown that there was no significant relationship between health expenditures and service coverage (maternal services, child services, and both of them). Table 2 shows the result of the regression fit.

**Figure3:**
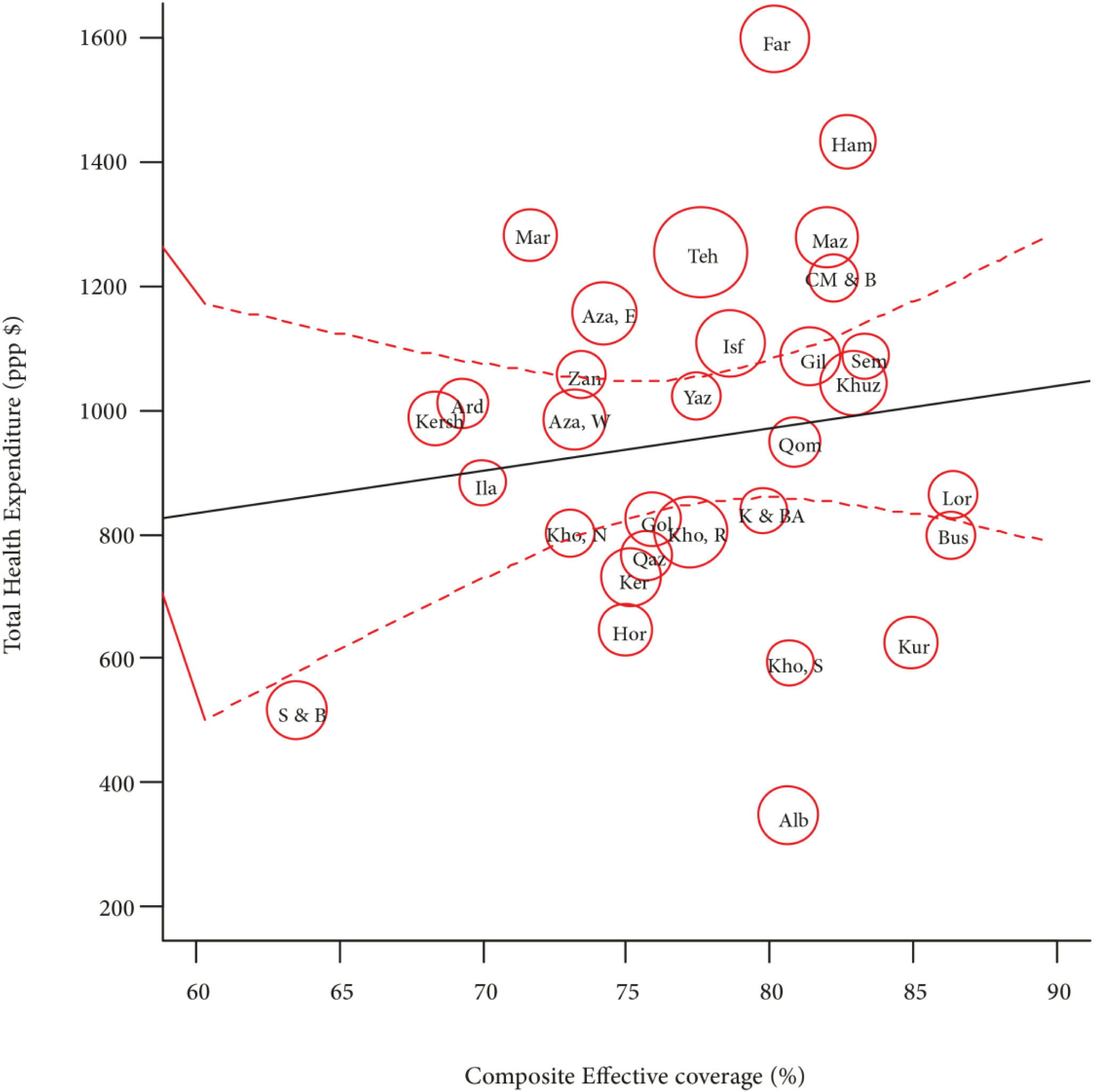
Relationship between composite effective coverage (six Interventions) by provinces and Total. Health Expenditure per capita

**Table 2:**
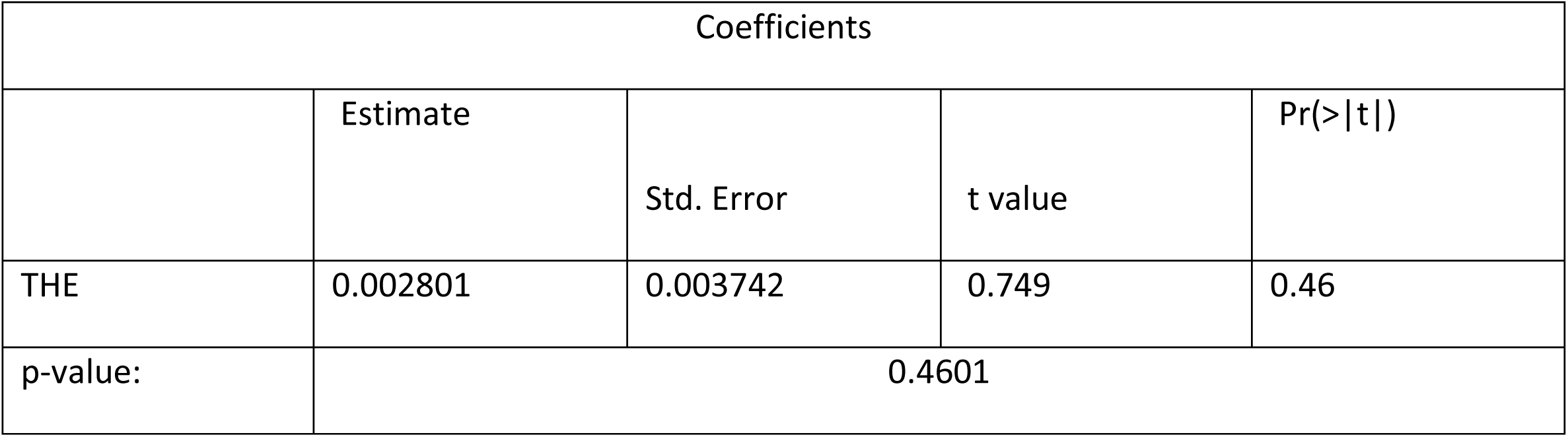
Regression fit between health expenditure per capita and the effective services coverage of PHC services in 2010.

| Coefficients |  |  |  |  |
| --- | --- | --- | --- | --- |
|  | Estimate | Std. Error | t value | Pr(>\|t\|) |
| THE | 0.002801 | 0.003742 | 0.749 | 0.46 |
| p-value: | 0.4601 |  |  |  |

## Discussion

This study aimed to estimate both crude and effective coverage of key maternal and child primary health care services at the provincial level as well as the impact of health expenditures per capita on the coverage. Our results show that there was a gap between crude and effective coverage of PHC indicators. Moreover, it was found that, maternal health interventions have higher coverage than children’s health interventions in the provinces. Although health expenditures had no significant relationship with effective service coverage, the provinces with the lowest coverage had also the lowest per capita health expenditures. The effective coverage values varied from 33.3% for care of children with diarrhea in Kermanshah province, to 100% for ANC, DTP3, and screening for hypertension and diabetes during pregnancy in some provinces. It is noteworthy that, a low effective coverage of key maternal and child health services indicates that individuals are not receiving the maximum possible health gain from existing health services (20).

The services reviewed in this study are provided free of charge in the public sector of Iran and, as mentioned earlier, they have a long history (before the Alma-Ata Declaration) worldwide (5). It is possible to receive these services in Iran in state-own facilities free of charge, even without having any insurance; however, it is also possible to receive these services from the private sector, and if the private facility has a contract with public health insurance funds, the provided service would be covered by insurance. Moreover, these services are completely provided by the public sector in the rural areas[21,22]. The indicators measured in this study are the main parts of the PHC interventions for improving maternal and child health. The results indicate that there is still a long way to reach the desired coverage and achieving UHC at the national level[20]. Even though these services have always been more available in rural areas, the coverage of all the included indicators still is lower than those of urban areas, which needs more investigations to clarify its causes. In Iran, the copayment for rural insurance scheme that receive health services from the public sector is around 5%. So, in order to use specialized services, people with rural insurance need to be formally referred to specialized centers by healthcare providers, to be able to use the copayment. However, there is no such requirement for people living in cities[21]. Although the PHC system is much older in rural areas of Iran compared to the cities, it seems that the quality of services provided in these centers is not acceptable by rural residents; therefore, the coverage of services provided for them is lower. Moreover, it is also necessary to examine the patient pathways in rural areas. The results of this study show that the location of the received PHC services (public/private) should be asked in future surveys, and then their satisfaction level *with the quality of* care that they receive should be examined.

In this study, we measured the geographic distribution of a range of PHC services coverage. In recent years, there is a general tendency to use a set of tracer indicators with targets for a selected set of health services’ priorities to evaluate the performance of health systems worldwide[23]. After introducing UHC as the main target of SDGs, the WHO has introduced the CCI (composite coverage index) as a service coverage index, which measures countries’ performances[18]. Accordingly, similar studies performed in Mexico[24], Bangladesh[25], and worldwide[10] have evaluated the performance of their health systems using a set of composite indicators[24].

In this paper, 14 out of 31 provinces performed well in providing PHC services. Four provinces are located above the third quarter zone, which had an excellent performance. However, in per capita expenditure estimation, these four provinces did not necessarily have high per capita costs. Hence, it seems that their optimal performances were not associated with a higher per capita expenditure. In Sistan-Baluchestan province with the most unfavorable indicators, per capita expenditures was lower compared to the other provinces of the country. Although no significant relationship was found between health care coverage and per capita health expenditure, it seems that the allocation of resources to low-income provinces must be revised.

In recent years, HTP, was implemented on accelerating progress to UHC, which also had several aims and one of them was to improve PHC services. The source of information used in this study was DHS, and no other similar household survey has been conducted since 2010. Therefore, it is not possible to assess the impact of HTP actions on crude and effective service coverage indicators. So, it is necessary to design and implement a similar survey to assess the current performance of PHC in Iran. By doing this, it is possible to evaluate the effect of the HTP and Iran’s position on reaching the UHC[9].

Services for the prevention and treatment of NCDs have been integrated into Iran PHC system, and their risk factors were screened among pregnant women[26]. The results of this study show that Iran’s PHC system has an acceptable performance in terms of screening NCDs in this population. However, to assess the performance of the health system in the fields of preventing and controlling NCDs, it is necessary to perform this assessment on the general population.

Geographic variations in coverage of maternal and child health indicators can reveal the existing inequities between and within countries[27]. In this regard, the lowest maternal indicators were seen in Sistan-Baluchestan province. This province is known as one of the most deprived provinces in the country[27]. Therefore, it is necessary to tailor interventions in each province according to their population needs, rather than performing a similar plan for all provinces. In fact, in Iran, there is a centralized approach to plan and implement PHC interventions[28]. Recently, WHO has developed PHCMI (Primary Health Care Measurement and Improvement) initiative, which is also implemented in Iran. The initiative aims to assess the current status of PHC services within countries, and then to use that assessment to inform strategies for the improvement of PHC at the local level. Hence, it is expected that the improvement at the sub-national levels would occur; therefore, it is recommended to measure effective coverage indicators after the implementation of the plan in the country[29].

The maternal mortality rate is decreasing in Iran, and according to the latest available report[11], it has a ratio of 16 per 100,000 live births, which has had an appropriate decrease compared to the rate in 2010[29]. This decrease implies that the measures taken to improve maternal care in Iran have been successful[26]. Notably, vaccination coverage is relatively high in Iran. Also, a high immunization coverage in children has been one of the major achievements of the health system in Iran for many years[30], and recently, it has succeeded to receive a certificate in the elimination and eradication of some infectious diseases (including polio[31,32], measles, and rubella[33]). Vaccination in Iran is performed free of charge in all state-owned health centers. In addition, DTP3 vaccine has been recently replaced by the DTP-Hepatitis B-Haemophilus influenza type b (Hib)[34], which should be considered in future assessments. In Iran, there is still no population-level indicator collection to assess the effective vaccination coverage, so this should be included in future plans.

The coverage of care of diarrheal and acute respiratory infection disease is lower than the other studied indicators. Although indicators related to care for infectious disease among children are far from 100% coverage, the children mortality rate (under fiveyears old) has a declining trend since 2010 (15.3 to 9.4). However, children mortality might be caused by these diseases, which should be considered in future assessments[11,26].

The share of PHC from THE based on the national health accounts at the whole country was 4.9% in 2010, and this was about 4.7 % in 2017. Although THE per capita in 2017 has increased by about 26 percent compared to 2010, the share of PHC services of THE has slightly changed, which may provide challenges for PHC expansion initiatives[35]. On the other hand, the WHO has recently recommended that the share of PHC in the public budget should be as much enough to ensure that countries move towards UHC. Accordingly, in Iran, this share is 47.9 %, while it is estimated to be less than 40% at the global level[36]. In this study, the provincial estimations of PHC’s share of THE have not been performed, and it is recommended that these estimations should be made in Iran at the provincial level, as well.

## Conclusion

There is a need to focus on both the quality of the PHC services and reaching them to achieve the goals of UHC. The concept of effective coverage has not been incorporated into health system performance assessment in Iran, yet. In addition, there is a need to measure the quality of data to gauge the strengths and weaknesses of the county’s primary health care system. Also, it is recommended to assess PHC performance based on some recently proposed frameworks such as PHCMI. Notably, Iran has a top down centralized PHC system. The results show that, although the average PHC performance is almost appropriate and some of the surveyed areas have very high performances, some low-income and deprived provinces experience substantial challenges. It is recommended that PHC plans should be formulated across the country and then implemented according to the local health needs and priorities. Moreover, it is recommended that policymakers pay greater attention to bolstering health service quality, particularly for services that have achieved a relatively high utilization, to optimize population health outcomes. Furthermore, regarding the fact that PHC services does not have a good share of THE, so there is a need to allocate additional public funds to PHC network. The next issue that should be considered is to strengthen the quality of essential *primary health care services provided for children*, because children indicators are far from reaching the intended target. Finally, it can be concluded that PHC system should be strengthened in Iran, as an essential step toward achieving universal health coverage by considering national and sub-national needs and priorities.

## Data Availability

The datasets for the current study are available from the corresponding author on reasonable request.

## List of abbreviations

ANC: Antenatal Care
CCI: Composite Coverage Index
DHS: Demographic and Health Survey
DTP-3: Diphtheria-Tetanus-Pertussis_3
HTP: Health Transformation Plan
MICS: Multiple Indicator Cluster Survey
NCDs: Non-communicable Diseases
PHC: Primary Health Care
PHCMI: Primary Health Care Measurement and Improvement
SDGs: Sustainable Development Goals
THE: Total Health Expenditure
UHC: <u>Universal Health Coverage</u>
WHO: World Health Organization

## Acknowledgements

The authors thank the National of Health Research(NIHR), Iran for their support and for free access to the original data of demographic and health survey.

## Author Contributions

Conceptualization: EAh, ZHA. Data curation: ZHA, EAb. Formal analysis: EAh, EAb. Funding acquisition: None. Methodology: EAh, EAb. Project administration: EAh. Writing – original draft: EAh, EAb. Writing – review & editing: ZHA, EAh, EAb,SS,BH.

## Corresponding author

Correspondence to Elham Ahmadnezhad.

## Ethics approval

This article is the secondary analysis and no need for ethical approves.

## Consent for publication

Not applicable.

